# Inherent Bias in Electronic Health Records: A Scoping Review of Sources of Bias

**DOI:** 10.1101/2024.04.09.24305594

**Authors:** Oriel Perets, Emanuela Stagno, Eyal Ben Yehuda, Megan McNichol, Leo Anthony Celi, Nadav Rappoport, Matilda Dorotic

## Abstract

1

**Objectives:** Biases inherent in electronic health records (EHRs), and therefore in medical artificial intelligence (AI) models may significantly exacerbate health inequities and challenge the adoption of ethical and responsible AI in healthcare. Biases arise from multiple sources, some of which are not as documented in the literature. Biases are encoded in how the data has been collected and labeled, by implicit and unconscious biases of clinicians, or by the tools used for data processing. These biases and their encoding in healthcare records undermine the reliability of such data and bias clinical judgments and medical outcomes. Moreover, when healthcare records are used to build data-driven solutions, the biases are further exacerbated, resulting in systems that perpetuate biases and induce healthcare disparities. This literature scoping review aims to categorize the main sources of biases inherent in EHRs.

**Methods:** We queried PubMed and Web of Science on January 19th, 2023, for peer-reviewed sources in English, published between 2016 and 2023, using the PRISMA approach to stepwise scoping of the literature. To select the papers that empirically analyze bias in EHR, from the initial yield of 430 papers, 27 duplicates were removed, and 403 studies were screened for eligibility. 196 articles were removed after the title and abstract screening, and 96 articles were excluded after the full-text review resulting in a final selection of 116 articles.

**Results:** Systematic categorizations of diverse sources of bias are scarce in the literature, while the effects of separate studies are often convoluted and methodologically contestable. Our categorization of published empirical evidence identified the six main sources of bias: a) bias arising from past *clinical trials*; b) *data-related biases* arising from missing, incomplete information or poor labeling of data; *human-related bias* induced by c) implicit clinician bias, d) referral and admission bias; e) diagnosis or risk disparities bias and finally, (f) biases in machinery and algorithms.

**Conclusions:** Machine learning and data-driven solutions can potentially transform healthcare delivery, but not without limitations. The core inputs in the systems (data and human factors) currently contain several sources of bias that are poorly documented and analyzed for remedies. The current evidence heavily focuses on data-related biases, while other sources are less often analyzed or anecdotal. However, these different sources of biases add to one another exponentially. Therefore, to understand the issues holistically we need to explore these diverse sources of bias. While racial biases in EHR have been often documented, other sources of biases have been less frequently investigated and documented (e.g. gender-related biases, sexual orientation discrimination, socially induced biases, and implicit, often unconscious, human-related cognitive biases). Moreover, some existing studies lack causal evidence, illustrating the different prevalences of disease across groups, which does not *per se* prove the causality. Our review shows that data-, human- and machine biases are prevalent in healthcare and they significantly impact healthcare outcomes and judgments and exacerbate disparities and differential treatment. Understanding how diverse biases affect AI systems and recommendations is critical. We suggest that researchers and medical personnel should develop safeguards and adopt data-driven solutions with a “bias-in-mind” approach. More empirical evidence is needed to tease out the effects of different sources of bias on health outcomes.

**CCS Concepts:** • **Computing methodologies** → **Machine learning**; **Machine learning approaches**; • **Applied computing** → **Health care information systems**; **Health informatics**; • **Social and professional topics** → **Personal health records**; **Medical records**.

**ACM Reference Format:** Oriel Perets, Emanuela Stagno, Eyal Ben Yehuda, Megan McNichol, Leo Anthony Celi, Nadav Rappoport, and Matilda Dorotic. 2024. Inherent Bias in Electronic Health Records: A Scoping Review of Sources of Bias. 1, 1 (April 2024), 24 pages. https://doi.org/XXXXXXX.XXXXXXX

## 2 INTRODUCTION

In the United States, The Health Information Technology for Economic and Clinical Health Act [66], enacted in 2009, fueled innovation programs that promoted the use of EHRs in creating artificial intelligence (AI) solutions. Globally, similar programs have promoted the use of EHRs in algorithms and systems that aim to improve the operational and administrative efficiency of healthcare systems. The exponential growth of patient data has opened up promising avenues for enhancing how physicians take care of their patients. Only in 2020, around 2.31 exabytes of new data were generated in healthcare [124]. To improve efficiency in analyzing this vast amount of information and deriving actionable insights to enhance decision-making, AI is used in multiple domains, including healthcare. AI systems are trained on data collected in medical facilities, mainly EHRs which contain patients’ medical history. EHRs are typically maintained by a health provider over time and may include all of the key administrative clinical data relevant to that person’s care, including demographics, progress notes, problems, medications, vital signs, past medical history, immunizations, laboratory data, and radiology reports [69].

The training of AI on EHRs creates opportunities for improved efficiency in analyzing the vast amount of health data and for deriving actionable insights to enhance decision-making at a significantly lower cost. The benefits include early prediction of disease risks, forecasting prognosis and development of disease, identifying to-human-eye-hidden data patterns, and providing personalized treatment recommendations for optimizing the delivery of treatment to large amounts of patients (which would not be feasible otherwise due to the limited availability of human resources). For example, AI can significantly more accurately than human experts screen and detect the risk of dementia [13] and improve the prediction of long-term recurrence risks for patients with ischemic cerebrovascular events after discharge from the hospital [143]. Moreover, the adoption of AI solutions to classify skin lesions using images is superior to humans in the challenging task of diagnosing the fine-grained variability in the appearance of skin lesions [41]. AI solutions can also be used to diagnose diseases such as diabetic retinopathy in parts of the world where there are too few ophthalmologists to read the fundus photographs and to follow up with each diabetic patient [144].

However, while powerful, AI solutions are only as good at making judgments as the data they are based on and humans that interpret and oversee its results [120]. Consequently, models based on the evaluation of EHRs can be subject to different types of biases [16]. In this paper, we define the biases inherent in EHRs or medical AI as the presence of systematic errors, unintentional prejudices, and inaccuracies in medical data that can affect the quality and fairness of healthcare delivery and decision-making [54]. EHR may hold inherent biases that occur through how data is collected, analyzed, or used. These inherent biases may be hidden in the data as they have many different sources, which may be hard to identify or correct, but they are likely to exacerbate the biases that the users of such data make (AI, health systems, or health professionals). For example, EHRs can be subject to biases arising from collecting, reporting, labeling, or designing EHRs, biases arising from implicit, unconscious stereotypes that healthcare professionals or algorithm engineers have as humans [45] or inherent biases in algorithms and machine learning (ML) models. These diverse biases can produce systematic errors in the data used to train AI solutions, which in turn spread through the system and exacerbate biased decision-making of healthcare professionals. As such, these biases limit reliability, perpetuate disparities, and increase health inequities when developing data-driven solutions from biased EHRs to treat patients. Nevertheless, it is unclear from which sources biases arise and how different types of biases specifically affect healthcare outcomes. Finding potential remedies for the biases requires a deep understanding of its causes, which currently only surfaces in the literature [60].

The outcomes of biases are potentially grave. For example, disease misclassification occurs as a consequence of bias hindering machine learning models developed on EHRs [94]. A potential source of bias emerges from disparities in diagnosis and prognosis for patients with different demographic or societal/environmental attributes. In a study of all reported trials supporting FDA oncology drug approvals between 2008 and 2018, Black and Hispanic races are consistently underrepresented compared with their burden of cancer incidence [87]. Previous research has shown disparities in diagnosis between patients with different types of cancers [57] or differences in risk of HIV between Black and White men when accounting for confounding variables [57]. The use of such data to build models that predict the health needs based on the existing patterns in the EHRs would be consequently skewed in favor of overrepresented rather than underrepresented populations [105].

On top of more known sources of inherent bias in data, the literature also mentions less investigated sources such as the biases connected to the usage of medical devices/algorithms or implicit (and often unconscious) human biases when treating patients with different demographic characteristics [45]. These biases are sometimes perceived to originate from the measurement error of the machinery, but at other times they are attributed rather to the differences in diagnoses or prognoses those devices create for patients with different demographic attributes. For example, significant variability was found in measurements of oxygen saturation levels for a given SpO2 level in Black patients relative to Hispanic, Asian, and White patients due to differences in how devices function on patients with different skin characteristics [140]. Although patients with or without hidden hypoxemia had similar clinical and demographic attributes, those with hidden hypoxemia had more comorbidities [68]. On the other hand, some studies showed that biased outcomes may also arise due to implicit, unconscious human biases of healthcare professionals (due to stereotypes, prejudice, or other unconscious cognitive biases), which consequently negatively affect the quality of care [45]. Such biases may affect the referrals to treatment. For example, previous studies have shown a systematically lower percentage of preventive cervical cancer screening for lesbian/gay patients than for bisexual and heterosexual patients across five US federally qualified health centers [60].

From the current findings, it is unclear how biases differ across diverse sources, what their idiosyncrasies are, and how to address these various biases. It remains unclear what exactly causes various biases and how they affect the decision-making of health professionals and AI systems. It does not help that the terminology related to biases is vast and varied such that different types of biases are confused in terminology, while biases coming from different sources are treated as synonyms, which further increases confusion. The identification of biases in the data used to train AI is a key pillar for constructing fair and responsible AI to be used in decision-making. Therefore, identification of the sources of biases is an important step in developing solutions to address these biases. Indeed, in health care, growing attention is given to understanding responsible approaches to the development, implementation, management, and governance of AI [120].

This project aims to categorize the main sources of inherent biases in EHRs documented in the existing literature. In particular, the purpose of this scoping review is to answer the following questions:

1. *What is the extent and nature of inherent bias in electronic health records collected in medical facilities?*
2. *What are the prominent sources of bias in EHRs?*

A scoping review methodology was used [129] for a comprehensive overview of the existing literature on these topics.

## 3 METHOD

This review was conducted and reported following the Preferred Reporting Items for Systematic Reviews and Meta-analyses extension for Scoping Reviews (PRISMA-ScR) [120].

### 3.1 Protocol Registration

A protocol for this scoping review has been registered with OSF. The protocol is available at: https://osf.io/skdm7

### 3.2 Eligibility Criteria

The eligibility criteria for the selection of papers included in our systematic review of the literature were as follows: (1) articles must explore some type of bias that may affect EHRs, (2) be written in English, (3) published between 2016 and 2023 (since the major investigative work on AI biases started in 2016), (4) published in peer-reviewed journals, and (5) include empirical evidence of some form of bias. We included most study designs, from experimental and quasi-experimental study designs to randomized controlled trials, non-randomized controlled trials, before and after studies, interrupted time-series studies, analytical observational studies, and descriptive observational studies. We excluded opinion pieces, conceptual papers, and medical case studies as these can’t indicate a larger-scale phenomenon or statistically show the existence of bias. To narrow the search and identify the sources of bias in classic medicine (e.g., medical care administered in a hospital or medical facility) articles were also excluded if they focused on a subject out of scope for this review, including psychiatry, psychology, dentistry, veterinary, and medical education.

### 3.3 Search Strategy

We applied the search strategy to two large databases, PubMed [22] and Web of Science, as their scope and coverage of the topics at hand are relevant and extensive. The initial query consisted of several key subject terms including and adjacent to: “Bias” in combination with terms such as “electronic health record,” “EHR,” “healthcare provider,” “clinical dataset,” “disparities,” “racial,” “ethnic,” “implicit,” “gender,” “unconscious,” “socioeconomic,” “machine” etc. The queries aimed to cover diverse sources of bias that may be inherent in EHRs and the medical field. The final search strings used for each database are presented in Appendix A. These queries yielded roughly 8,000 search results. Hence, we iteratively added conjunctions and exclusions of topics out of scope for this review. After the first analysis of the title and abstract, articles discussing disparities in medical education, workplace or wage disparities of medical practitioners, and gender-related bias in surgical residency were excluded. We narrowed the search strategy years to 2016-2023 to decrease the number of results while providing a time-relevant overview of the literature, justified by the increased interest and resources invested in equitable care and fairness in machine learning in recent years [55]. In the next stages, while reviewing selected studies, we used the snowball sampling technique to identify relevant studies for our analysis that were referenced in included articles but which we missed in our initial search.

### 3.4 Study/Source of Evidence Selection

Based on the database query from the previous section, 410 articles were retrieved and uploaded to the “Covidence” systematic review management platform for the review process. Key sources were identified through discussion, from which minimal cross-referencing was conducted by the authors, resulting in 20 additional sources manually added to Covidence. To provide unified criteria for the inclusion of sources, based on the readings and discussion among authors in our interdisciplinary team, we have defined broad categories of bias as those related to data, humans, and algorithms or machinery. Of the 430 sources included for review, 27 duplicates were identified and removed, and 403 studies were screened for eligibility in two stages. First, titles and abstracts were screened for further analysis, with the main criteria being that the study analyzes some form of inherent bias (as identified earlier) in a medical context. This phase excluded 191 articles (Figure 1). Next, a full-text review was conducted on the remaining 212 articles. Sources were excluded based on the following criteria: 1. The study was not peer-reviewed; 2. Editorials, conceptual pieces without data, or a qualitative study design (21); 3. The topic was irrelevant to this study (6); 4. The study does not illustrate an inherent bias (24); and 5. The study shows prevalence or differences in groups of patients but it does not provide evidence of bias (45). All excluded papers were labeled by at least two reviewers, and any conflicts were discussed between reviewers and resolved by a third reviewer. This phase resulted in the exclusion of 96 articles. We note that the main reason why relatively many of the initial hits were excluded refers to the fact that articles did not test empirically for bias. This points out the deficit of empirical studies, despite the wide discussion of these issues. Eventually, 116 articles were included for the scoping review. The full inclusion process is displayed in Figure 1.

**Fig. 1.**
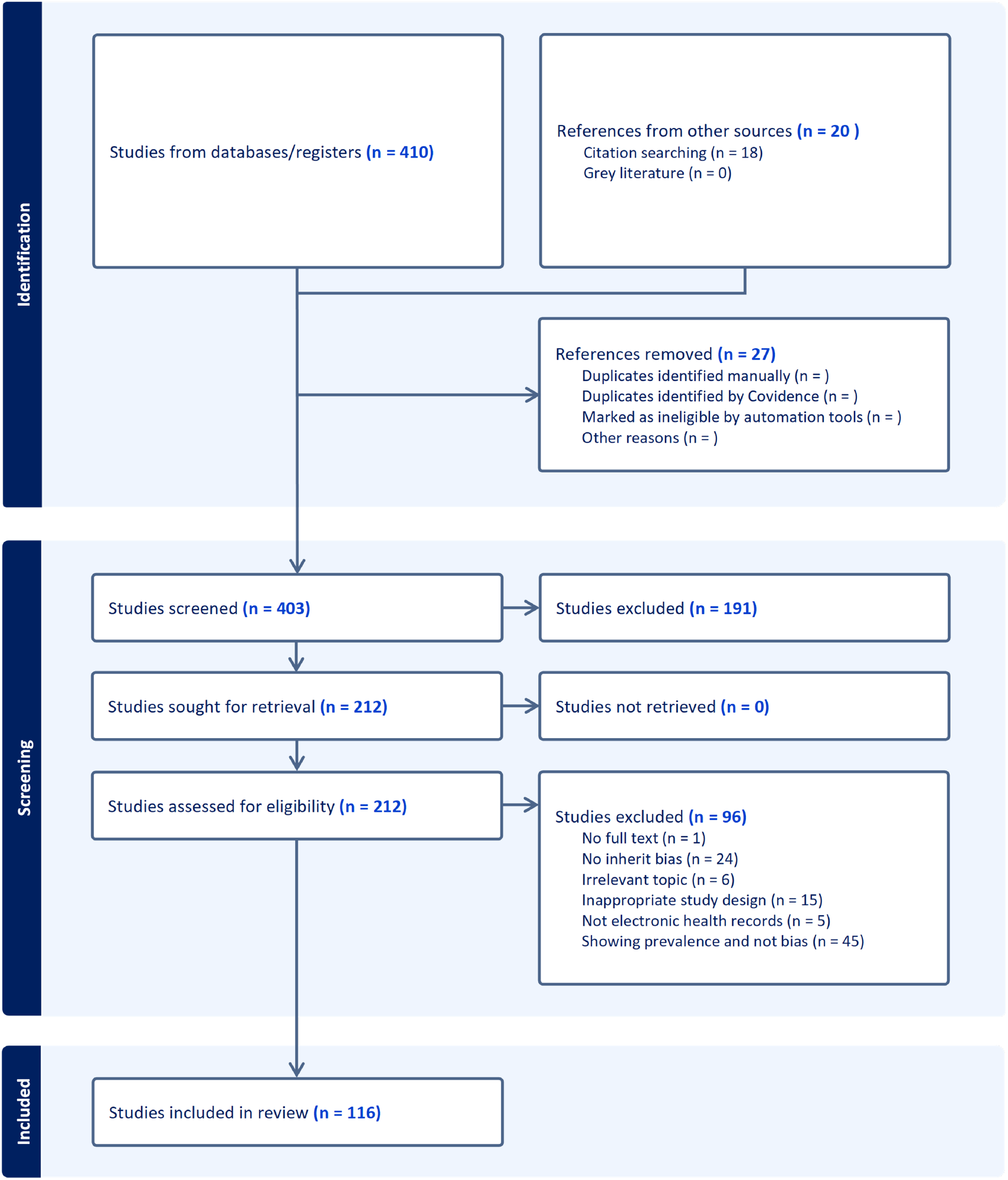
PRiSMA exclusion flowchart: the flow chart describes the number of filtered articles and the reasons for exclusion

### 3.5 Data Extraction/Data Items

An analysis of the articles included after applying the eligibility criteria and conducting a full-text review revealed that they could be grouped into six categories of sources of bias. These sources were further categorized into primary and secondary bias categories, and specific details about the bias characteristics, context, study methods, and findings relevant to the review question were extracted. This process was carried out independently by two or more reviewers and is presented in the synthesis of the results section.

## 4 FINDINGS

### 4.1 Important terminology issues identified in the studies

The above analysis of studies offered important insights. We found that the terminology of the phenomena varies across disciplines, areas, and studies. While the term “inherent bias” is prominent in medicine, “machine bias” and its synonyms are used in engineering and computer science literature, and “systemic, algorithmic, or AI bias” are terms prominently used in social science literature. This indicates the need for a careful and broad selection of keywords and mechanisms when reviewing the phenomena.

### 4.2 Important methodological issues identified in the studies

The most prominent methodological issue we found across studies is the problem that many studies, that use the term bias or indicate they study biases in healthcare, often do not employ the methodological approaches suitable for testing the causal effects that would show the impact or presence of a bias. As indicated above, most excluded studies (and related discussions among reviewers) concern studies showing the prevalence of a condition in different populations or other differences between groups. While biases certainly may arise from these differences, they do not unequivocally prove the bias itself or the cause of the bias. Merely showing differences in two populations (e.g., group A being more prone to an illness or less willing to receive treatment relative to group B) does not directly confirm inherent bias in EHRs or machinery, as there is no link established between the effect and bias and its outcomes. We illustrate the issue with a famous example that showed using the overall (past) spending on healthcare as a proxy for predicting (future) healthcare needs/likelihood to develop complications results in machine bias that prioritizes treatment for affluent White patients at the cost of less represented Black patients, who were less represented in the data due to their lower utilization of the healthcare system [105]. Showing the resulting causal effect of using such an unrepresentative proxy which would lead to a biased algorithm, that would in turn give biased predictions for underrepresented groups, is an example of proving inherent bias.

## 5 SOURCES OF BIAS IN EHRS

Through an in-depth analysis of the included 116 papers, co-authors of this study classified the papers into six main sources of bias: (a) clinical trial-induced biases, (b) missing, incomplete, or poor labeling biases; (c) implicit clinician biases, (d) referral and admission biases, (e) diagnosis, prognosis or risk disparities biases, and (f) medical machinery (AI) biases. It is noteworthy to point out that diverse other names exist for these types of bias, which we reference in the text when discussing those biases. The prevalence of each type of bias among the selected 116 papers is presented in Table 1. We note that these different sources of biases are not mutually exclusive; they may overlap, lead to one another, or be otherwise interrelated. We have used these six categories holistically to show that biases differ in their sources, and these differences should be considered, both by researchers and practitioners.

**Table 1.** Types of bias and number of detected and screened publications.

| Type | Form of Bias | n | Prevalence | References |
| --- | --- | --- | --- | --- |
| Data-related Biases | Clinical Trial Induced Bias | 5 | 4% | [12, 19, 40, 87, 109] |
|  | Missing, Incomplete, Poor Logging of Information | 14 | 12% | [3, 21, 31, 39, 54, 67, 73, 81, 84, 100, 103, 104, 108, 119, 126] |
| Human-induced Biases | Implicit Clinician Bias | 19 | 16% | [7, 24, 29, 35, 36, 43, 45, 75, 76, 86, 110, 112, 122, 130, 131] |
|  | Referral and Admission Bias | 41 | 35% | [6, 10, 15, 18, 32–34, 37, 42, 43, 46, 47, 52, 59, 60, 64, 70, 72, 77–80, 82, 85, 89–93, 97, 99, 107, 113, 115, 118, 121, 123, 138, 139, 141, 142, 145] |
|  | Diagnosis, Prognosis or Risk Disparities Bias | 30 | 26% | [1, 8, 20, 23, 25, 30, 38, 49–51, 57, 61, 62, 65, 74, 83, 88, 95, 100, 117, 125, 132, 134, 135, 137] |
| Machine Bias | Medical Machinery Bias | 9 | 8% | [4, 9, 11, 68, 106, 116, 127, 133] |

### 5.1 Clinical trial-induced bias

Clinical trials stand as pivotal elements in medical research, crucial for the evaluation and validation of new therapeutic interventions, drugs, and standards of care. These studies systematically assess the impacts of specific interventions on human subjects by juxtaposing them with control groups, either untreated or given a placebo (i.e., a treatment without therapeutic effects investigated in the treatment, e.g. harmless pill) [114]. The ethical and logistical challenges inherent in clinical trial recruitment necessitate not only enlisting a sufficient number of participants to ensure statistical power [58] but also striving for a sample that mirrors the diversity of the target population [114]. This stage poses an ethical question to the study’s conductor since the goal is to recruit an adequate number of participants to generate enough data to answer the research questions at hand, but it is usually impossible to choose a completely random sample group and at the same time representative of the general population or even the of the study’s population [48]. This endeavor is crucial for minimizing *selection bias*, a form of bias that occurs when the trial population does not adequately represent the broader community, potentially skewing the trial’s outcomes and applicability [63]. The selection bias is closely related and sometimes equated to the concepts of *underrepresentation bias* or *informative presence bias*. The informative presence bias occurs in instances when those who have poorer health have more data recorded than healthier patients or in other similar instances when one patient group is represented more in the sample than other groups but this composition does not reflect the proportions in the general population of the studied disease[63]. As a result, employed AI solutions would prioritize patients in better-represented groups and make significant errors in the interpretation and prediction of the health needs of the underrepresented population, thereby further inducing health disparities between these groups. In the identified literature, the underrepresentation bias was shown to affect the decision thresholds, administration of care, or the medical procedure conducted in the study with no other plausible source of bias affecting the decision.

The under- and over-representation biases based on conclusions from previously conducted clinical trials come both from the self-selection in scientific publishing as well as the disparities in the socio-economic systems that define access to healthcare around the world. Publication bias, for instance, emerges from the tendency to publish studies with positive outcomes more frequently than those with negative or inconclusive results, leading to a skewed perception of an intervention’s efficacy [96]. Another critical concern is the underrepresentation of certain demographic groups, either due to their scarcity in the general population or the presence of systemic barriers that deter participation and thereby create missingness in the dataset. Such disparities can introduce “class imbalances,” particularly detrimental in studies of rare diseases or for small communities with a certain sensitive feature on which AI bases prediction. This characteristic can significantly bias the development and evaluation of data-driven health solutions [128]. For example, AI models trained on datasets predominantly comprising data from one racial group are found not to perform accurately when applied to underrepresented groups [14]. The unrepresentativeness may occur due to the small samples (e.g. in minority populations), which may lead to underestimation or overestimation of risks in the subpopulation relative to the majority producing inaccurate or misleading recommendations. Deep learning algorithms used to predict acute kidney injury in adults significantly underperformed for female patients which represented just 6.4% of the data in the training dataset [126]. Similarly, the breast cancer survival rate algorithm underperformed for Black women which represented only 7.8% of the data [30]. Moreover, systematic reviews have highlighted significant gaps in the inclusion of racial and ethnic information in clinical research, underscoring the challenge of achieving representativeness [2]. Out of 38 studies that met the criteria of inclusion in the meta-analysis, 28 did include racial or ethnographic information (73%), but only 17 studies (45%) included information on Hispanic participation while comprising a mere 11.6% of participants out of the general population tested [109]. A database study of all reported trials supporting FDA oncology drug approvals granted between July 2008 and June 2018 showed that only 63% of trials reported at least one race and 36.9% had no mention of race. In 7.8% of trials that reported on all four races (White, Asian, Black, or Hispanic), only White patients’ representation matched their proportion based on cancer incidence and mortality in the United States, while Black and Hispanic populations were consistently underrepresented, and Asian overrepresented [87]. The disproportionality between clinical trial enrollment of certain ethnic minority groups and the prevalence of diseases among them is demonstrated empirically [12]. Less than 5% of African Americans participated in Diabetes mellitus clinical trials and only 32% took part in Antihypertensive and Lipid-Lowering Treatment to Prevent Heart Attack Trial [12]. Participation in medication and therapy programs in the U.S. also indicated racial disparities, as patients with Alzheimer’s disease and related dementias (ADRD) and diabetes were tested for their enrollment in the Medicare Medication Therapy Management (MTM) program. Significant disparities between Whites and Blacks were found within all study cohorts, as Blacks had significantly lower odds of enrolling in MTM programs compared to Whites [20]. Gender disparities are evident in the underrepresentation of women in cardiovascular trials. Only 33% to 38% of all clinical trials conducted between 2010-2020 consisted of women [12]. The lower participation rates of women in trials for hypertension, diabetes, hyperlipidemia, coronary artery disease, heart failure, and arrhythmia in recent years contradicted the prevalence of these diseases in women.

The repercussions of underrepresentation extend beyond the accuracy of clinical predictions to affect treatment paradigms and health outcomes across different population segments. Disparities in trial participation rates can lead to the development of treatment standards that do not adequately address the needs of all groups, particularly those historically marginalized or with limited access to healthcare resources [87]. Even more, the informative presence bias does not occur only between subjects, but also within-subject data when analyzing longitudinal patterns in the development of the disease. For example, longitudinal analyses of biomarkers are likely to show bias in hazard ratio estimates if biomarker values are analyzed predominantly based on informative visits (initiated by a patient), rather than noninformative visits (like routine check-ups). [56]. Even when both informative and noninformative visits are combined, the informative bias in biomarkers longitudinal analysis will form if the biomarkers are more volatile over time [63]. Addressing these challenges requires concerted efforts to enhance the inclusivity and equity of clinical research, ensuring that the benefits of medical advancements are accessible to all segments of society.

### 5.2 Missing, incomplete information and poor logging of information

Missing or incomplete information and poor logging of information refer to bias arising from the quality of electronic health records used for research. EHRs are susceptible to biases stemming from missing, incomplete, or inaccurately logged information. These inaccuracies can arise from various factors such as medical staff oversight, technical issues in logging data, poor labeling, patient non-disclosure, or systemic constraints. This review identifies studies addressing the prevalence and implications of such data deficiencies.

Besides the fact that some of the subgroups in a population may intrinsically have smaller samples in the population, other important reasons for underrepresentation in databases may have intrinsically social causes which may lead to the lower representation of some groups in the databases due to the missing information. Examples include the underrepresentation of groups based on some structural barriers like the socioeconomic lack of access (for example, to medical care) or subpopulation’s self-imposed exclusion due to the distrust in the system that may drive under-representation or other issues in self-reporting. Disparities in cancer diagnosis are found to be explainable by health insurance coverage, such that racial/ethnic minority children and adolescents are more likely to have an advanced cancer diagnosis compared with non-Hispanic Whites, which may relate to the lack of consistent healthcare access [136]. Neighborhood socioeconomic status (SES), rather than family SES, explained around 29% of racial disparities in the average rate of change in blood pressure trajectories for Black American versus non-Hispanic White preterm children [49]. Healthcare access due to social reasons, device allocation that differs between rural and urban areas or between hospitals, and measurement frequency which can be affected by the patient/doctor ratios or the location of the hospital, can all influence which patients will be included in the databases and with which likelihood [26].

In the realm of EHRs, a significant bias source is the inconsistent or erroneous recording of information, which can be seen as the labeling bias. For instance, discrepancies are found in the coding of uveitide ICD10 across the two most prominent EHR systems (Epic and MDIntelleSys) yielding different codes in 13 out of 27 diseases, leading to potential data fragmentation and analysis impact [108]. Moreover, multiple codes were used to describe more than one specific disease. Therefore, combining data from different EHR systems can lead to fragmented and incomplete data and may affect downstream analysis and conclusion. Moreover, socioeconomic data is often unrecorded, and analyses based solely on complete cases introduce considerable selection bias [81]. This bias was evident in research on epithelial ovarian cancer, which indicated that missing data is not at random since the cases with missing data correlated with poorer survival outcomes [126]. Biases arising from missing or poor quality data are also likely to have led to disparities in healthcare services to LGBTQ patients [60]. Missingness and labeling biases can become amplified over time or in subsequent iterations of the data. Biases in EHR can skew large-scale studies, as illustrated by a study examining the time-varying biases in genome-wide association studies, which could lead to inaccurate gene-phenotype associations [119]. The off-label use of open databases is another concern, potentially generating biased machine learning models due to overlooked bias in data collection or processing, a phenomenon dubbed “data crime” [119]. Therefore, researchers who look to incorporate two or more datasets in model development to increase robustness and generalizability ultimately may introduce this form of bias into their model.

Data preparation adds another layer of complexity, introducing biases through labeling, coding, and the use of proxies for social categories. For example, differences in data labeling practices between Europe and the USA can significantly affect AI applications in pathology [102]. In the US, it is little known that some European staining solutions include saffron dye for improved fibrous tissue delineation, which may not even be indicated in the data description, since such information may be assumed as given [102]. Often different labeling terminology and standards are employed across countries, inducing variation and confusion in the use of terms and abbreviations. This issue extends to variations in clinical note-taking and terminology, which influence machine learning predictions in critical care mortality and psychiatric readmissions [29]. Labeling biases and poor logging of information are likely to occur in numerous instances as medical professionals typically log information post-hoc and perceive these tasks as an administrative burden.

Even when data is labeled correctly, it is often not advisable to use some features or models that must use proxies of variables for the information that is missing. The use of proxies to circumvent the direct use of sensitive attributes like race, gender, and ethnicity in health predictions introduces measurement biases. A seminal study demonstrated that using healthcare costs as a proxy for healthcare needs led to a biased prioritization of affluent White patients over Black American patients with comparable disease burdens [105]. This example underscores the need for careful consideration in data handling and model development to prevent perpetuating existing disparities.

### 5.3 Implicit Human Biases

In addition to data-related biases, healthcare practices and Electronic Health Records (EHRs) are influenced by the inherent, often unconscious biases of health professionals. These implicit biases entail the subconscious stereotypes healthcare providers hold, leading to skewed assessments of patient needs and medical conditions based on sensitive attributes such as race, gender, socioeconomic status, and medical history. These biases, which are typically involuntary and irrational, tend to be related to the implicit beliefs of healthcare professionals about the natural differences between genders, races, or ethnical minorities [45].

Healthcare professionals, like the general population, harbor implicit biases that reflect societal stereotypes about race, gender, and ethnicity. Such biases, inadvertently recorded in EHRs, contribute to perpetuating healthcare inequities. The training of medical students, which emphasizes pattern recognition and stereotypical symptoms, may further embed these biases. Consequently, implicit bias adversely impacts diagnostic and treatment decisions, thereby exacerbating healthcare disparities. For instance, disparities in diagnosing malingering—where patients are unjustly accused of exaggerating medical issues—disproportionately affect men and Black patients [132]. Encoding such diagnosing in EHR has serious consequences on future medical care, the likelihood of admittance and receiving treatment.

Implicit biases have tangible consequences on medical care, particularly for marginalized groups such as ethnic minorities, immigrants, economically disadvantaged individuals, sexual minorities, and others, leading to systemic under-treatment and diagnostic inaccuracies. These biases manifest in various healthcare aspects, including initial disease diagnosis, and the timeliness and accuracy of diagnoses and treatments across several medical domains like neurodegenerative, pulmonary, cardiac diseases, and oncology [16, 84]. Implicit bias has been shown to lead to substantive physician errors in predicting heart attack such that physicians overtest predictably low-risk patients and under test predictably high-risk patients because they predominantly rely on simple heuristics like chest pain [101]. An AI model that learns from the resulting data will inevitably be biased in the same way.

Research also indicates that implicit biases contribute to significant physician errors, such as the misallocation of diagnostic resources based on simplistic heuristics rather than patient risk levels [122]. Patient reluctance to disclose information about prescribed medications to clinicians may signal mistrust stemming from previous experiences or the healthcare provider’s perceived judgment about the patient’s treatment adherence [29, 86].

### 5.4 Referral and admission bias (including access to care)

Biases related to clinician referrals and patient admissions significantly influence healthcare access and research participation. An admission bias arises when the selection criteria for research participation or medical care disproportionately exclude individuals based on discriminatory characteristics, leading to skewed study populations or unequal access to care [43]. Similarly, referral bias occurs due to systematic differences in who gets referred for further evaluation or treatment, often disadvantaging certain groups [33]. An example may be seen in studies that showed that minorities with worse objective dry eye parameters received fewer prescription treatments or procedures than White patients [33].

This review focused on identifying studies that highlighted disparities in access to care and referral patterns across different treatments, such as cancer prognosis, cardiovascular diseases, and appendicitis, based on sociodemographic factors rather than objective medical criteria. These barriers often stem from implicit biases and socioeconomic constraints, leading to delayed or foregone care [64, 124]. The impact of racial and socioeconomic status on access to care is well-documented, with minority groups and those from lower socioeconomic backgrounds facing significant barriers to receiving treatment and diagnoses at earlier, more treatable disease stages [14, 18]. Lower socioeconomic strata patients were diagnosed at a later stage of anal cancer disease, had worse survival than wealthier patients, and were less likely to receive radiation therapy [23].

Furthermore, studies have shown that disparities in care access and referral patterns manifest early in life, with children from African American backgrounds experiencing lower admission rates and follow-up care in intensive settings [59]. Gender disparities also contribute to these biases, affecting referral rates to specialists and hospital admissions [5]. For instance, young women are less likely to undergo certain procedures like percutaneous coronary intervention compared to their male counterparts, resulting in higher in-hospital adverse event rates [109].

Socioeconomic status further compounds these issues, with individuals from lower socioeconomic strata receiving suboptimal care for conditions such as blood pressure management [44]. These systematic biases not only impact patient outcomes but also influence the representation of diverse populations in clinical data, perpetuating biases in data-driven healthcare solutions.

In summary, implicit biases related to clinician referrals and patient admissions create significant barriers to equitable healthcare access and treatment. Addressing these biases requires concerted efforts to ensure that healthcare and research practices are inclusive and representative of the diverse populations they serve.

### 5.5 Diagnosis, prognosis, or risk disparities bias

In addition to the biases in admission and referral to further treatment, implicit biases also occur at the later stages of the patient’s journey. Disparities in the diagnosis, prognosis, and risk assessment within medical datasets emerge as a critical form of inherent bias, significantly influenced by patient demographics such as gender, SES, and race. A notable body of research highlights these disparities, particularly in the context of cancer diagnosis and treatment initiation stages. For instance, disparities based on SES and racial groups are evident in the diagnosis and treatment of various cancers [23, 132, 136]. Specifically, racial and ethnic disparities have been documented in the delayed diagnosis of appendicitis among Black children, where delays were associated with prior emergency department visits and the absence of imaging during these visits [59].

Moreover, disparities in cancer diagnosis have been linked to differences in health insurance coverage, leading to advanced cancer diagnoses among racial/ethnic minority children and adolescents compared to their non-Hispanic White counterparts, attributed to inconsistent healthcare access [136]. Similarly, patients from lower socioeconomic backgrounds are diagnosed at later stages of diseases such as anal cancer, experiencing poorer outcomes and reduced access to treatments like radiation therapy [23]. The influence of neighborhood SES, as opposed to individual or family SES, accounts for a significant portion of the racial disparities observed in health outcomes, such as blood pressure trajectories among preterm children [49].

Access to healthcare and the allocation of medical devices, which varies between rural and urban locations or among different hospitals, along with disparities in the frequency of medical assessments, can dictate the inclusion of patients in medical databases, further perpetuating these biases [44].

Additionally, conditions such as insulin resistance have been shown to contribute to racial disparities in breast cancer prognosis, highlighting the complex interplay between medical conditions and demographic factors [50]. Heightened HIV risk is found among Black men compared to White men, underscoring the role of specific risk factors unique to different racial groups [57].

This review identified thirty studies showcasing this category of bias, marking it as the most prevalent type of bias encountered. Such findings underscore the urgent need for strategies to mitigate these disparities, ensuring equitable healthcare outcomes across all patient demographics.

### 5.6 Data bias in medical devices and algorithms

Medical machinery bias stems from discrepancies in how algorithms are programmed and the calibration of devices, potentially leading to measurement inaccuracies for patients with diverse ethnic, physiological, and biological characteristics. This bias may arise from technical errors in medical equipment [106, 116, 127, 133] or from underlying differences in measurements due to factors not related to the patient’s medical condition. Within medical facilities, patients undergo frequent automatic testing using equipment such as pulse oximeters, EKGs, ECGs, and thermometers, alongside semi-automatic tests involving healthcare professionals (e.g., hematology analyzers, biochemistry analyzers, scales, and blood glucose monitors). The outcomes of these assessments play a crucial role in patient evaluation and treatment planning.

We noted nine articles in this review highlighting this form of bias. Specifically, pulse oximeters exhibit reduced accuracy in patients with darker skin pigmentation, an issue attributed to device miscalibration and the lack of diversity in development phases [106, 116, 127, 133]. Similar disparities in device accuracy affecting measurements like oxygen saturation, body temperature, and blood pressure [44] have been documented, often resulting from insufficiently diverse calibration populations [26]. A systematic review of mechanical ventilation studies found that AI applied to mechanical ventilation has limited external validation and model calibration, with a substantial risk of bias, significant gaps in reporting, and poor code and data availability [53]

Such discrepancies in device performance can introduce biases into clinical data, potentially influencing treatment decisions, such as the administration of supplemental oxygen or the preference for certain temperature measurement methods, thereby affecting diagnoses and treatments for specific racial subgroups. For example, studies have identified an increased risk of occult hypoxemia in Black patients compared to White patients with identical oxygen saturation measurements [116, 133].

Additionally, benign ethnic neutropenia (BEN), a condition where individuals (commonly of African, Middle Eastern, and West Indian descent) present with neutropenia (absolute neutrophil count < 1500/µL) without an increased infection risk, exemplifies bias stemming from physiological variances across different patient groups [4, 9]. The biased interpretation of such measurements or conditions perpetuates sub-optimal care for specific patient demographics, further encoded into EHR systems, highlighting the critical need to address medical machinery bias to ensure equitable healthcare delivery. In addition, substantial biases in AI algorithms arise due to implicit human biases of programmers who develop algorithms, similar to the implicit biases outlined for health professionals. It is well documented that experts in computer science and AI developers are a fairly homogeneous group with the mental mindset of a Western White male [17].

## 6 DISCUSSION

### 6.1 How diverse sources of biases can perpetuate one another

In the preceding discussions, we have endeavored to delineate and exemplify various biases originating from distinct sources. Adopting a process-oriented perspective, we aim to elucidate how these diverse sources of biases can interact and potentiate each other. Figure 2 conceptualizes a typical patient journey, highlighting how inherent biases might emerge at various junctures and proliferate over time within a technocratic framework that leverages data-driven AI systems in healthcare. This depiction underscores the cumulative nature of biases, illustrating their potential to compound and exacerbate disparities in patient care and outcomes within AI-enhanced healthcare environments.

**Fig. 2.**
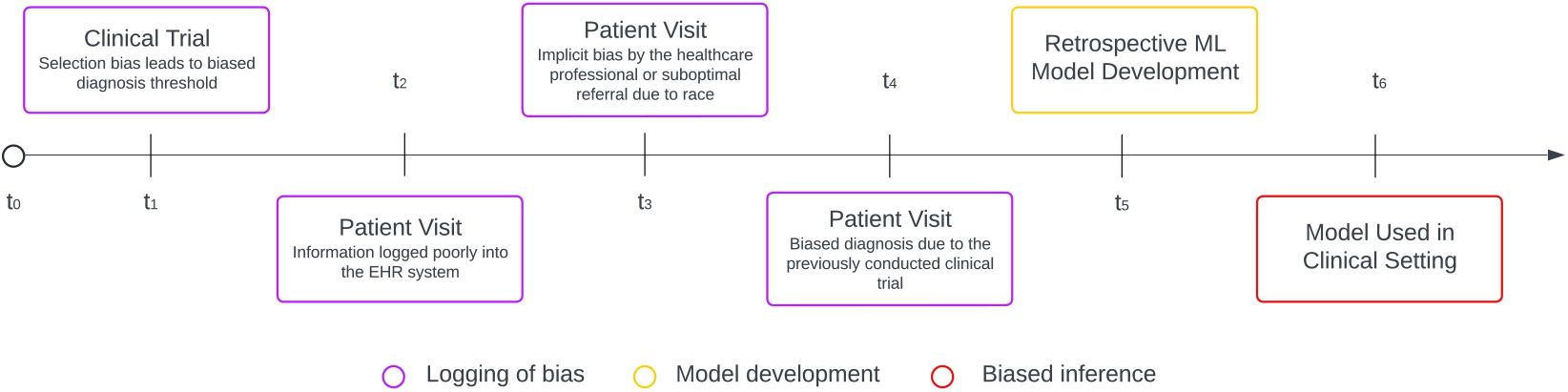
Illustration of the source of bias on a timeline. A patient’s timeline from inception. *t*_0_ represents a clinical trial that results in a biased diagnosis threshold towards specific patient populations, and *t*_1_ − *t*_4_ represents the lifetime of a patient in the healthcare system until a particular time when the patient receives biased care due to implicit bias, or bias in medical machinery. Incomplete information and biased clinical decisions are logged into the EHR system. At time *t*_5_, a retrospective ML model is developed based on previously collected data. Which consequently results in biased inference, perpetuating the bias toward other patients

Consider a hypothetical patient, Anne, at the initial point in time, *t*_0_, who begins experiencing symptoms necessitating a medical evaluation and possibly treatment. The primary step involves comparing her symptoms and medical history against existing knowledge and clinical trial data. Depending on Anne’s ethnicity, gender, or socioeconomic status, she might find that the algorithms governing admission prioritize her case differently compared to other patients with similar symptoms. In the second stage, while waiting for admission, Anne might struggle with self-administered questionnaires, especially questions about her socioeconomic status and sexual orientation, due to their complexity or her reluctance to provide comprehensive responses. Objecting to these questions on moral grounds and privacy concerns, she decides to omit some answers and fabric some others. Subsequently, Anne meets with a healthcare professional who, despite their best intentions, appears significantly different from her in many respects, making it difficult for her to feel understood. This professional relies on an AI healthcare system that seems unable to fully “understand” Anne. The system, concurring with the doctor, diagnoses her in a manner that suggests she does not require hospital admission or further treatment. Left uncertain about her actual health risk despite her symptoms, Anne wonders if the system simply fails to recognize her unique needs. She is discharged, and the AI model, employing machine learning algorithms, records her case to refine future treatment predictions for similar cases. At the end of the shift, the doctor writes a very short note about Anne’s case in the records. Despite Anne’s experience, the hospital lauds the model for its operational efficiency.

### 6.2 Discussion of evidence

While this review aims to categorize and point out possible sources of hidden biases in EHRs, we emphasize this review serves only as a “call to action” and is far from covering the entire spectrum of biases currently residing in EHRs and medical data in general. This scoping review synthesizes the documented sources of bias in the current literature on EHRs, incorporating 116 articles that met the inclusion criteria. The review introduces inherent or hidden bias within EHR data, which poses significant challenges to developing machine learning models and the generalizability and precision of research findings. The inherent bias in EHRs can significantly affect retrospective studies and algorithmic model development. This review proposes a categorization of inherent bias into six distinct categories based on the source of bias: implicit clinician bias, bias in medical machinery, disparities in diagnosis, prognosis or risk, referral and admission bias, biases resulting from clinical trials, and issues related to missing, incomplete, or poorly logged information. Increased discussion about potential sources of biases in healthcare has increased awareness, but diverse sources of bias identified in this review are still scarcely accounted for in research and development of AI solutions in healthcare.

The presence of these biases was consistent across various settings, populations, and EHR systems, indicating a broad applicability of this categorization framework. Notably, the review highlights the frequent misidentification or underreporting of bias types in the literature, where terms like disparities, equity, or differences in risk or prevalence in disease diagnosis often mask inherent bias.

The methodology of this review, grounded in clinical and human-machine interaction insights followed by a targeted literature search, has led to a focused and informative collection of articles. Referral and admission bias emerged as the most frequently documented category, suggesting that disparities in healthcare access—driven by factors like race, gender, and SES precede and influence the care process, thus introducing a selection bias in the development of machine learning models. The prominence of referral and admission bias reporting is understandable given that the objective data for analyzing this bias is more readily available. However, this finding also warns about the importance of providing empirical evidence on other sources of bias and the importance that transparency of databases and access to open sources have in the mitigation of AI biases.

The second most prevalent source of bias identified involves disparities in diagnosis, prognosis, or risk, highlighting how different risk factors and demographics can affect patient outcomes across various diseases, notably in oncology [23, 132]

Despite these challenges, advancements in artificial intelligence offer potential mitigations for human-induced biases. For instance, AI models have shown promise in addressing biases in radiographic measures and in recognizing the physical causes of pain across racial and socioeconomic groups, thereby accounting for a larger proportion of disparities in pain than traditional measures. For example, relative to standard measures of pain severity graded by radiologists, which accounted for only 9% of racial disparities in pain, AI predictions accounted for 43% of disparities, with similar results for lower-income and less-educated patients which stem from the racial and socioeconomic diversity of the training set [27].

This review underscores the critical need for awareness and corrective measures against inherent biases in EHR data to improve the fairness and effectiveness of machine learning applications in healthcare.

Addressing racial and other disparities due to data incompleteness is crucial for enhancing equity in health systems. Protocols and guidelines for routinely collecting and reporting disaggregated ethnicity data are recommended [16]. Initiatives like The HL7 Gravity project, which seeks to standardize social determinants of health measures for interoperable electronic health information exchange, are pivotal [98]. Synthetic data approaches in machine learning also play a vital role in compensating for the lack of diverse annotated medical data, ensuring analytical processes reflect the general population’s diversity [28, 111].

Implicit clinician or healthcare provider bias represents a significant challenge, impacting ground truth (GT) for retrospective models and hindering the ability of models to eliminate existing biases. Addressing this requires educational interventions to increase awareness among medical students and practitioners [5]. Efforts to increase diversity within hospitals and medical schools may contribute to reducing such biases.

The literature indicates that machine or AI bias, resulting from historical biases in diagnoses, prognoses, and clinical decisions logged in EHRs, can be perpetuated by machine learning (ML) models. This underscores the necessity for critical evaluation of data and the acknowledgment of potential biases in medical machinery, such as the differential risks indicated by pulse oximeters for patients of different races with identical readings [127], and varied infection risks for patients with benign ethnic neutropenia (BEN) [9].

Furthermore, missing or poorly logged information in EHRs poses an additional source of inherent bias in EHR. This includes compatibility issues with ICD diagnosis codes across different EHR systems, which can introduce biases into ML models developed using data from multiple sources [108]. Researchers aiming to enhance model robustness and generalizability by integrating multiple datasets must navigate these challenges to avoid introducing biases.

### 6.3 Limitations

This scoping review is subject to several limitations that warrant careful consideration. Primarily, the inclusion of articles exclusively from PubMed and Web of Science introduces a potential database selection bias, potentially overlooking relevant research published in other languages or databases. Additionally, by focusing on peer-reviewed articles from 2016 to 2023, the review may omit earlier or more recent developments related to bias in electronic health records (EHRs). The exclusion of non-peer-reviewed literature could also contribute to publication bias, possibly exaggerating the prevalence of inherent bias, since the studies that show the bias may be more likely to get published than studies with inconclusive or nonsignificant results. Furthermore, the absence of a quality and rigor assessment for the included studies might impact the review’s reliability. Lastly, the generalizability of the findings is potentially limited, as the review may not encompass bias sources specific to certain demographics, healthcare environments, or EHR systems. Future research should aim to mitigate these limitations and provide a more comprehensive understanding of bias in EHRs.

### 6.4 Conclusions

We want to emphasize that we approach the conclusions with epistemic humility acknowledging that the current evidence only scratches the surface of the potential problems of AI implementation in healthcare globally and due to the centralization of evidence in Western societies we lack a holistic picture from those who have not gotten so far “a seat at the proverbial table”. This article aims to highlight these inequalities and call for more research into the sources of bias that permeate global health records and AI model developments. With that said, the integration of AI in healthcare presents a transformative potential for enhancing efficiency, enabling the analysis of vast datasets, and facilitating the extraction of actionable insights to inform decision-making at reduced costs. AI’s capabilities extend to more accurate screening and detection of dementia risks [14] and improve predictions of long-term recurrence risks for ischemic cerebrovascular events post-discharge [143]. Moreover, AI advancements contribute to the refinement of risk scoring and mortality prediction models for a variety of conditions [71]. However, the effectiveness of these advancements is contingent upon the availability of accurate and unbiased data, as biases present in Electronic Health Records (EHRs) can significantly impact diagnosis and treatment decisions. Contrary to the focus on machine bias in computer science literature, our review identifies six critical sources of “inherent biases” within EHRs that are less recognized and addressed in the medical field. These biases include (a) implicit clinician bias, (b) bias in medical machinery, (c) disparities in diagnosis or risk assessment, (d) referral and admission bias, (e) biases stemming from clinical trials, and (f) the presence of missing or incomplete information.

These challenges contribute to perpetuating disparities and biases in algorithmic solutions, affecting their fairness, outcomes, and the trust of professionals and the public place in AI technologies. Our findings add to the importance of addressing these inherent biases enhancing the effectiveness and adoption of data-driven solutions and AI in healthcare.

## Data Availability

A scoping review, all articles are available in the bibliography to this manuscript

## 7 ACKNOWLEDGMENTS

We express our sincere appreciation to the Big Biomedical Data Lab at BGU for their valuable input and discussions. Special thanks are also extended to Natalie Anne Jansen, M.D., and Alon Dagan, M.D., from the Beth Israel Deaconess Medical Center, for their insightful contributions to our discussions.

